# A Genomic Survey of SARS-CoV-2 Reveals Multiple Introductions into Northern California without a Predominant Lineage

**DOI:** 10.1101/2020.03.27.20044925

**Authors:** Xianding Deng, Wei Gu, Scot Federman, Louis du Plessis, Oliver G. Pybus, Nuno Faria, Candace Wang, Guixia Yu, Chao-Yang Pan, Hugo Guevara, Alicia Sotomayor-Gonzalez, Kelsey Zorn, Allan Gopez, Venice Servellita, Elaine Hsu, Steve Miller, Trevor Bedford, Alexander L. Greninger, Pavitra Roychoudhury, Lea M. Starita, Michael Famulare, Helen Y. Chu, Jay Shendure, Keith R. Jerome, Catie Anderson, Karthik Gangavarapu, Mark Zeller, Emily Spencer, Kristian G. Andersen, Duncan MacCannell, Clinton R. Paden, Yan Li, Jing Zhang, Suxiang Tong, Gregory Armstrong, Scott Morrow, Matthew Willis, Bela T. Matyas, Sundari Mase, Olivia Kasirye, Maggie Park, Curtis Chan, Alexander T. Yu, Shua J. Chai, Elsa Villarino, Brandon Bonin, Debra A. Wadford, Charles Y. Chiu

## Abstract

The COVID-19 pandemic caused by the novel coronavirus SARS-CoV-2 has spread globally, resulting in >300,000 reported cases worldwide as of March 21st, 2020. Here we investigate the genetic diversity and genomic epidemiology of SARS-CoV-2 in Northern California using samples from returning travelers, cruise ship passengers, and cases of community transmission with unclear infection sources. Virus genomes were sampled from 29 patients diagnosed with COVID-19 infection from Feb 3rd through Mar 15th. Phylogenetic analyses revealed at least 8 different SARS-CoV-2 lineages, suggesting multiple independent introductions of the virus into the state. Virus genomes from passengers on two consecutive excursions of the Grand Princess cruise ship clustered with those from an established epidemic in Washington State, including the WA1 genome representing the first reported case in the United States on January 19th. We also detected evidence for presumptive transmission of SARS-CoV-2 lineages from one community to another. These findings suggest that cryptic transmission of SARS-CoV-2 in Northern California to date is characterized by multiple transmission chains that originate via distinct introductions from international and interstate travel, rather than widespread community transmission of a single predominant lineage. Rapid testing and contact tracing, social distancing, and travel restrictions are measures that will help to slow SARS-CoV-2 spread in California and other regions of the USA.

## Introduction

The novel severe acute respiratory syndrome coronavirus 2 (SARS-CoV2), which causes coronavirus disease 2019 (COVID-19), is a pandemic that has infected more than 531,000 people around the world and caused more than 24,000 deaths as of March 26th, 2020 (1), including 85,500 cases in the United States and >3,500 in California. The exponential growth in the number of cases has overburdened clinical care facilities and threatens to overwhelm the medical workforce. These numbers underestimate the true number of infections due to the relative paucity of diagnostic testing to date (2) and the presence of asymptomatic or mild cases (3, 4). As a result, California, along with many other states and countries, has issued a “shelter-in-place” policy for all residents, effective March 20, 2020. These unprecedented measures have disrupted daily life significantly for ∼40 million inhabitants for an indefinite period, with the potential for profound economic losses (5).

Until late Feb 2020, the majority of infections identified in the United States were related to travelers returning from high-risk countries, repatriated citizens under quarantine, or close contacts of infected patients. Community spread, in which the source of the infection is unknown, has since been documented in multiple states. Furthermore, Washington State has reported a series of COVID-19 cases from Jan 21 to Mar 18, following the identification of an early case, WA1, on Jan 19, suggesting the presence of a persistent transmission chain in the community [Bedford, et al. 2020].

Genomic epidemiology of emerging viruses has proven to be a useful tool for outbreak investigation and for tracking virus evolution and spread (6, 7). During the Ebola virus disease epidemic of 2013-2016 in West Africa, genomic analyses established that the outbreak had a single zoonotic origin (8), that two major viral lineages were circulating (9), and that sexual transmission played a role in maintaining some transmission chains (10). Viral genome sequencing also uncovered the route that Zika virus traveled from northern Brazil (11) to other regions, including Central America and Mexico (12) and the Caribbean and United States (13). However, real-time genomic epidemiology data of COVID-19 to inform public health interventions in California have been lacking.

We recently developed a method called metagenomic sequencing with spiked primer enrichment (MSSPE) to rapidly enrich and assemble viral genomes directly from clinical samples (14). Here we used this method to recover viral genomes from COVID-19 patients in Northern California and perform phylogenetic analysis to better understand the genetic diversity of SARS-CoV-2 in the US and the nature of transmission of virus lineages in the community.

## Results

We generated virus genomes of SARS-CoV-2 lineages in California from nasal swab samples from 29 patients, available through the University of California San Francisco (UCSF) hospital, the Santa Clara County Public Health Department, and the California Department of Public Health **(Figure 1A and Table S2)**. Presumptive positive cases were confirmed to be SARS-CoV-2 positive by testing using a CDC assay approved by the FDA EUA (Emergency Use Authorization) on February 4th, 2020 *(14)*. Samples to be sequenced were selected for broad representation across different counties and association with known outbreaks. The 29 infected patients spanned 9 counties in Northern California **(Figure 1B)** and included (i) 9 samples airlifted from the Grand Princess cruise ship, during its excursions from San Francisco to Mexico and Hawaii in February and March 2020, (ii) 3 samples from a Solano County cluster, including the first reported case of community transmission in the United States, and (iii) 5 patients from a Santa Clara County cluster associated with workspace transmission. Fourteen of the 29 patients did not have a source of transmission that could be readily identified by travel history or epidemiological investigation.

**Figure 1.**
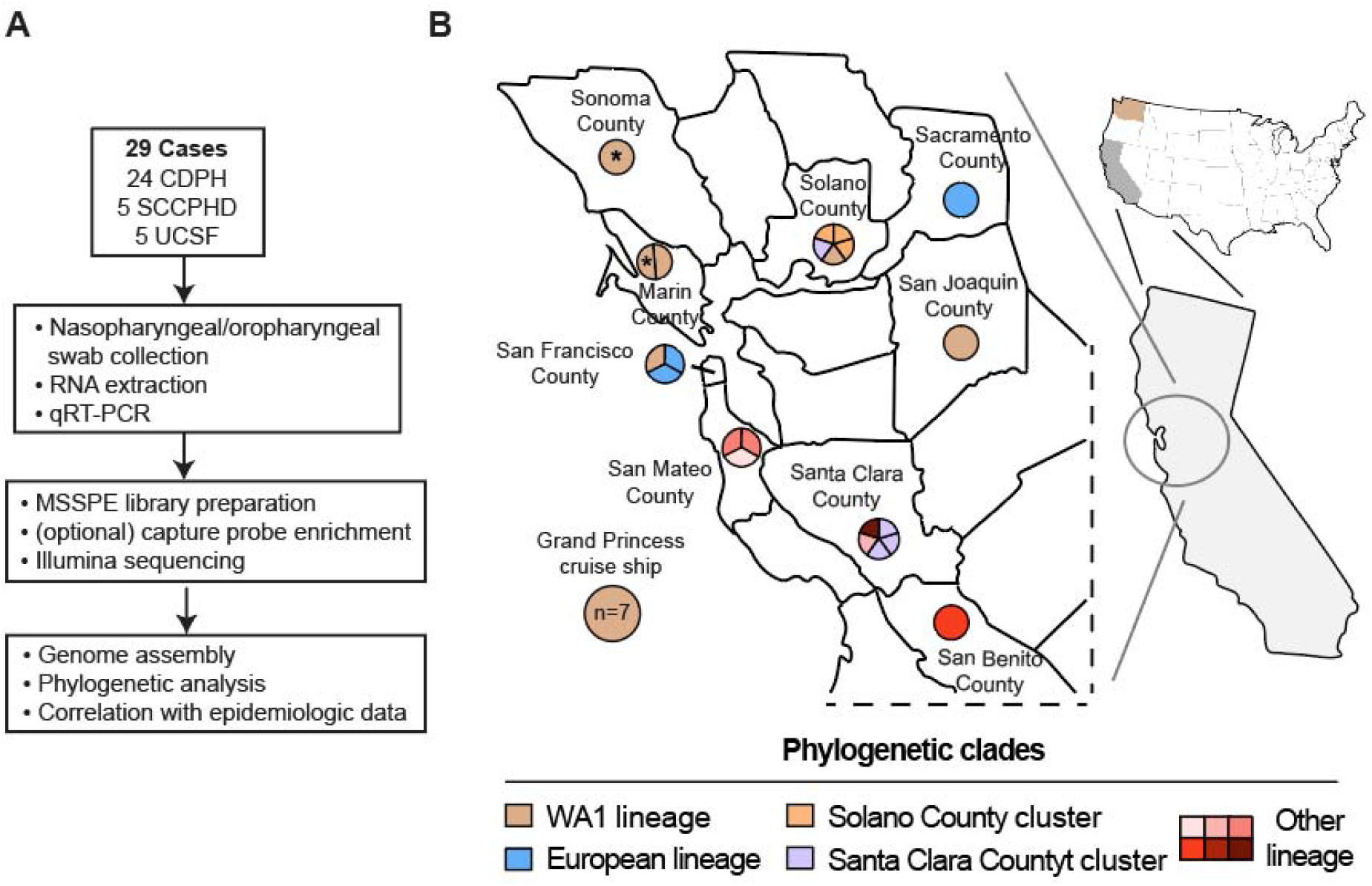
Genomic survey of SARS-CoV-2 genomes in Northern California. **(A)** Analysis workflow. **(B)** Map of the Northern California survey region. The pie charts for each county are subdivided according to the number of patients whose viral genome was sequenced, and the color corresponds to the viral lineage as determined by phylogenetic analysis. Passengers who were on the Grand Princess ship during the first cruise to Mexico are denoted by an asterisk.

We performed MSSPE on each sample to enrich for the SARS-CoV-2 RNA genome (15), followed by metagenomic next-generation sequencing (mNGS) of pooled and indexed samples on Illumina NextSeq or MiSeq instruments (16, 17) **(Figure 1A)**. Virus loads for the 29 patients ranged from 1.3⨯10^4^ - 2.5⨯10^8^ copies/mL. An average of 31 million reads (interquartile ratio, IQR, 23-57 million) were generated per sample, and virus genomes were assembled by mapping to reference genome NC_045512 (Wuhan-Hu-1). The assembly yielded 28 SARS-CoV-2 genomes with genome coverage >60%, and were thus included in this study. We also retained one partial genome (UC29) with 16.2% coverage, as a full genome from this individual had been already sequenced from a high-titer earlier sample by the US CDC (MT027062). The median coverage achieved across all samples was 96.5% (IQR 84.0%-99.7%).

Phylogenetic analysis shows that the SARS-CoV-2 genomes generated in this study are dispersed across the evolutionary tree of SARS-CoV-2 viruses, estimated from 342 genomes available on GISAID as of March 17th, 2020 **(Figure 2A)**. These viruses included 2 viruses likely related to lineages from Wuhan or other regions of China, 4 viruses related to those circulating in Europe, 11 viruses positioned in small independent clusters and not part of a larger clade, and 13 viruses associated with the Washington State (WA1) clade **(Figure 2B)**.

**Figure 2.**
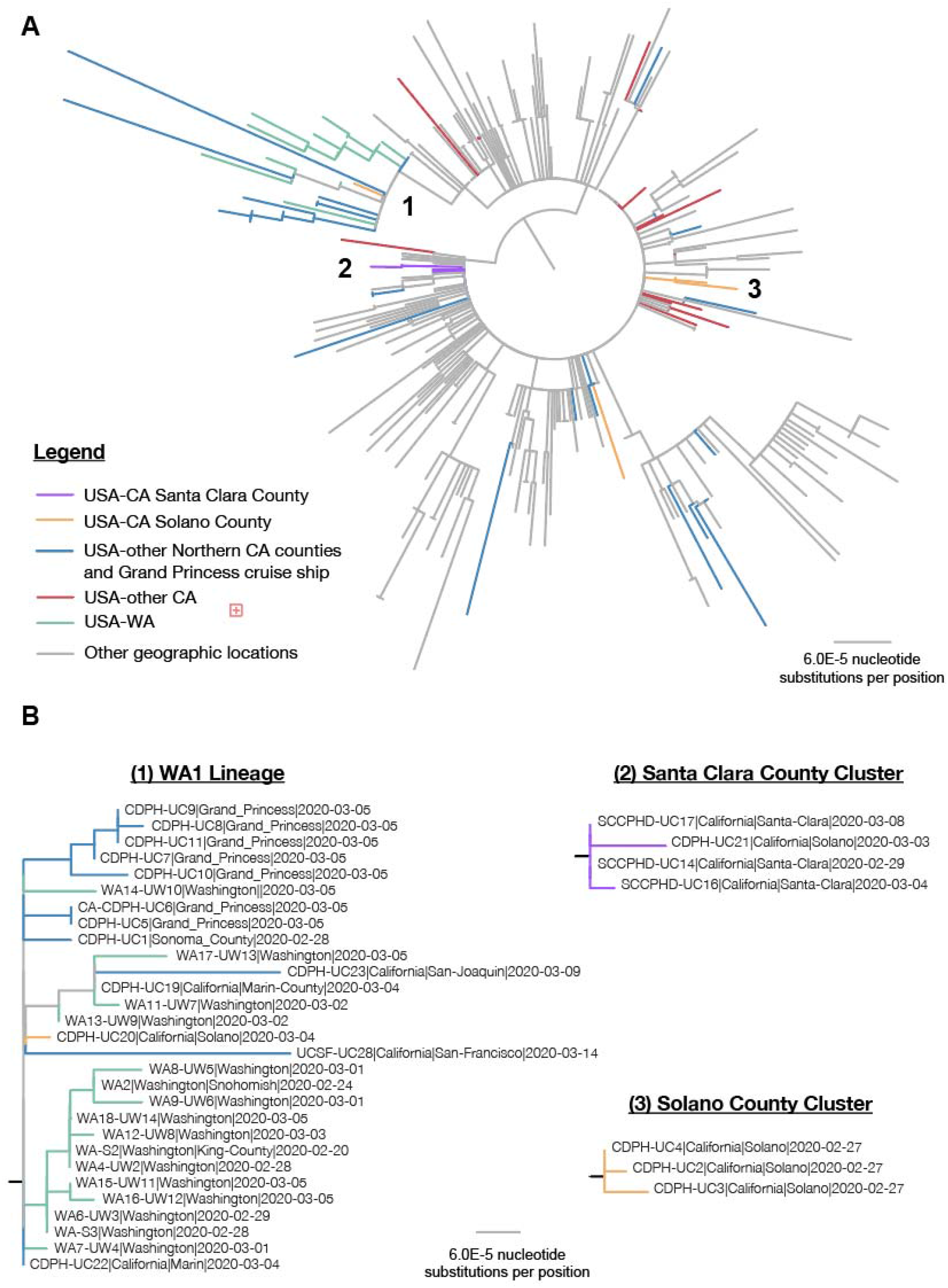
Phylogeny of SARS-CoV-2 lineages in California. **(A)** Phylogenetic tree of 342 SARS-CoV2 genomes along with the 29 genomes in this survey. Multiple sequence alignment of all available SARS-CoV-2 genomes in GISAID as of March 22nd, 2020, was performed using MAFFT *(24*), followed by tree constructions using PhyML *(25*). Viral lineages from California or Washington State are colored, while those outside of California and Washington are shown in gray. **(B)** A zoomed view of individual phylogenetic clusters corresponding to the WA1 lineage (1), Santa Clara County cluster (2), and Solano County cluster (3).

A large outbreak was associated with travel on the US cruise ship Grand Princess, which undertook two consecutive voyages from San Francisco (to Mexico on February 11 - 22, and to Hawaii on February 22-March 4) with much of the same crew and a shared subset of passengers. Notably, the 9 available sequenced genomes from crew members and passengers aboard the cruise ship (UC1, UC5 to UC11, and UC19) all clustered within the WA1 lineage.

The first reported case from the United States within this lineage (WA1) was reported January 19th (18) [Bedford, et al. 2020] and therefore substantially predates the excursions of the Grand Princess cruise ship. The SARS-CoV-2 sequences from cruise ship passengers and crew all shared two single nucleotide polymorphisms (SNVs), C17747T and A17858G, as well as the triad of SNVs common to all WA1 lineages (C8782T, C18060T, and T28144C) **(Figures 2 and 3)**. In addition, all of the passengers on the second cruise to Hawaii (UC 5-11) and UC1, who was on the first cruise to Mexico, carried at least 2 additional mutations not observed in UC19, who also was on the first cruise. This indicates that the virus from UC19 is most likely basal to the clade containing the cruise ship strains, and that COVID-19 infections associated with the first cruise were passed onto passengers and crew on the second cruise. The chronology and phylogeny of the cruise ship outbreak, along with the predominance of the WA1 lineage in Washington State but not outside of the US, also suggest that the virus on the Grand Princess likely came from Washington State, although it is still possible that the cases originated from a different region in which the WA1 strain is circulating. In addition to the passengers and crew members on the Grand Princess, virus genomes sampled from five cases of community transmission in different counties of the Bay area (UC19, UC20, UC22, UC23 and UC28) were also phylogenetically placed in the WA1 lineage, suggesting possible community circulation of that lineage in California as well as on the Grand Princess ship.

**Figure 3:**
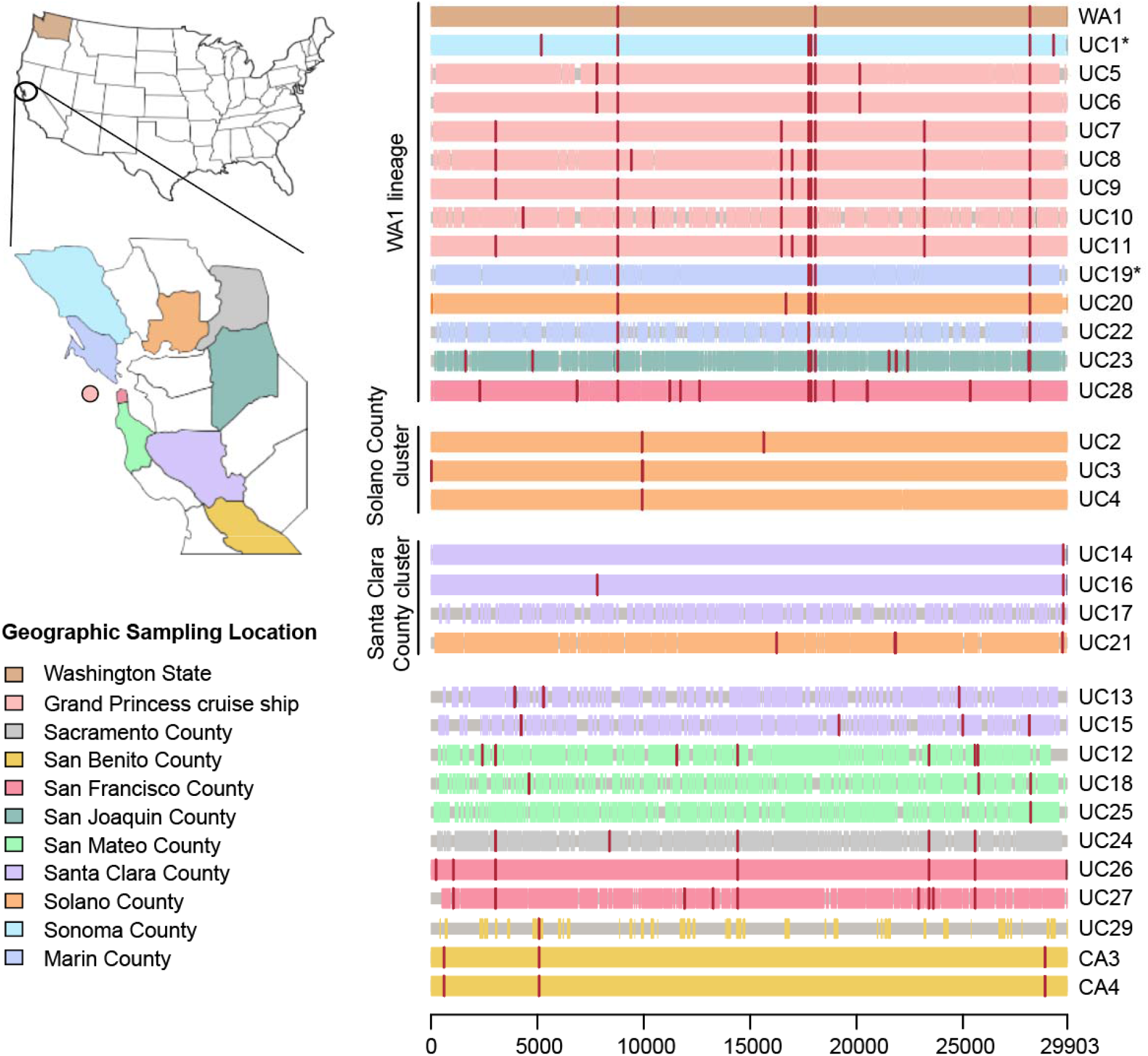
Multiple sequence alignment of all SARS-CoV-2 genomes reported across 9 counties and 1 cruise ship. Single nucleotide variants (SNVs) with respect to the reference strain (NC_045512) are shown in vertical red lines. Cases that are grouped into the WA1 cluster (defined as containing the C8782T, C18060T, T28144C SNVs) include the first identified case of COVID-19 infection (WA1) in the United States on January 15^th^, 2020, 9 passengers and crew members aboard the Grand Princess cruise ship, and 4 Northern California counties. Two additional SNVs, C17747T and A17858G, are common to SARS-CoV-2 viruses associated with the Grand Princess cruise ship. Single SNV variants C9924T and G29711T define groups of infected individuals in the Solano County (UC2, UC3, and UC4) and Santa Clara County (UC14, UC16, and UC17) clusters, respectively.

Two out of the 29 individuals examined in this study (UC12 and UC29) had COVID-19 infections associated with international travel or exposure to travelers. UC12 was a resident of San Mateo County with a travel history to Switzerland. The genome from UC12 fell within a large lineage containing many sequences from European residents or travelers from Europe and therefore it is likely the infection was acquired there. The sequence from individual UC29 (MT027062 is from the same patient) was a returning traveler from China and has an identical COVID-19 sequence to a household contact (MT027063). Their viral genomes were more closely related to ancestral lineages from China. Interestingly, three further genomes (UC24, UC26 and UC27) also grouped within a viral lineage with widespread circulation in Europe.

Individual UC27 was diagnosed after returning from a New York trip, raising the possibility of interstate transmission. Individual UC26 also reported domestic travel, while local resident UC24 had no known travel history, illustrating the existence of undetected virus transmission in the community.

A small cluster of 3 cases in Solano County includes the first reported instance of community transmission in the US on February 26 (UC4) **(Figures 2B and 3)**. The two other cases (UC2 and UC3) were healthcare workers who were taking care of patient UC4 and likely contracted the disease in the hospital, consistent with transmission of the disease from patient to health care providers (19). Two patients from San Mateo County (UC18 and UC25) were close household contacts (a husband and wife), and their viruses comprised a separate cluster with a virus from an infected person from China.

In Santa Clara County, cases with onset dates within two weeks of each other were associated with a large facility with multiple employers, large areas of shared space, and heavy pedestrian traffic. Out of five sequenced cases from this outbreak cluster (UC13 to UC17), three formed a distinct cluster by phylogenetic analysis (UC14 was an employee at the facility, while UC16 and UC17 were family members living in the same residence), indicating likely household transmission **(Figures 2B and 3)**. Notably, the genome from a Solano county resident (UC21) was also grouped into this cluster, suggesting possible circulation of a virus lineage between different counties. An active investigation regarding a putative epidemiologic link involving UC21 and individuals in the Santa Clara County cluster had been initiated prior to the genomic analyses and is ongoing at the time of publication. Two other workers (UC13 and UC15) also operated at the same building and had no direct contact with UC14; all three had different employers and no known social contact with each other. Consistent with this epidemiological data, the genomes (UC13-15) from the three workers in the large facility are distinct from each other (i.e. no identifiable shared SNV). Several possible scenarios could give rise to this observation, including the prospect that more than one virus lineage was introduced into the building via different events or the workers were infected as a result of community transmission.

## Discussion

The genomic epidemiology of the COVID-19 cases associated with community spread studied here do not show a predominant SARS-CoV-2 lineage circulating in Northern California. This epidemiological profile is distinct to that in Washington State, in which the vast majority of virus genomes sequenced to date belong to a single phylogenetic lineage, WA1 [Bedford, et al. 2020]. The WA1 lineage strains share common ancestry with an early imported lineage that seem to have established transmission in Washington State from January 2020. By contrast, in California, we conclude that multiple recent and unrelated introductions of COVID-19 into the state via different routes have occurred, giving rise to the diversity of virus lineages reported in this study, rather than cryptic transmission of a single pre-existing lineage among counties in California. We note that this does not exclude the possibility of cryptic transmission of multiple lineages in California, as the current level of sampling is not dense enough to estimate confidently the dates of the seeding events, nor the subsequent periods of cryptic transmission before a lineage was identified.

The co-clustering of the lineages from the Grand Princess cruise ship and those from several Northern California counties with community-transmitted cases in Washington State suggests an established lineage of SARS-CoV-2 in the USA. Notably, the WA1 lineage has been recently identified in COVID-19 cases in Illinois, Minnesota, Connecticut, and Utah. The early date and basal phylogenetic position of the WA1 lineage makes it likely that the direction of dissemination was from Washington State to other states; however that conclusion could change if further genomic sampling in the US revealed substantial levels of virus genetic diversity. SARS-CoV-2 mutates more slowly than many other human RNA viruses, on the order of 1 to 2 SNV mutations a month across its ∼29 kB genome (20), as it contains a nonstructural gene with proofreading activity (21). Despite this, the WA1 lineage of SARS-CoV-2 is readily identifiable with 3 key SNVs, C8782T, C18060T, and T28144C (Figure 3) which, at this stage in the epidemic, are distinct from other SARS-CoV-2 lineages.

Our epidemiological and genomic survey of SARS-CoV-2 has several limitations. First, this initial analysis represents relatively sparse sampling of cases. Undersampling of virus genomes is due in part to the high proportion of cases (80%) with asymptomatic or mild disease (3, 4, 20) and limited diagnostic testing for COVID-19 infection to date in California and throughout the United States. Second, the majority of samples analyzed were obtained from public health laboratories and thus may not be representative of the general population. Finally, phylogenetic clustering of viruses from different locations, such as Washington State and California in the same clade, does not prove directionality of spread. Despite this, our study shows that more robust insights into COVID-19 transmission are achievable if virus genomic diversity is combined and jointly interpreted with detailed epidemiological case data.

Public health containment measures such as prompt isolation and contact tracing, as performed in the Solano County and Santa Clara County clusters, become more difficult to maintain once a lineage becomes established in the community. Our data suggest concerning trends in this direction, such as the association between the WA1 lineage and community-acquired COVID-19 cases in several counties of Northern California, travel-associated introduction of SARS-CoV-2 into the San Francisco Bay Area from New York state, and a virus from the lineage associated with a Santa Clara County cluster detected in a resident of Solano County. Social distancing interventions, such as the “shelter-in-place” directive that was issued by the governor of California on March 20, 2020, may assist in stemming community spread of imported cases. If interstate dissemination of SARS-CoV-2 lineages within the US is common, then the suspension of non-essential domestic and international long-range travel may be necessary to prevent further importation of new cases in California and other states.

## Methods

### Ethics Statement

Clinical specimens were processed at the University of California San Francisco (UCSF) under protocols approved by the UCSF Institutional Review Board (protocol no. 10-01116, 11-05519). Specimens collected by the California Department of Public Health (CDPH) were de-identified and deemed not research or exempt by the Committee for the Protection of Human Subjects (Project number 2020-30) issued under the California Health and Human Services Agency’s Federal Wide Assurance #00000681 with the Office of Human Research Protections. A non-research determination for this project was also granted by Sonoma County as it was designated epidemic disease control activity, with collected data directly related to disease control.

### Sample Collection

Samples obtained from the UCSF Clinical Microbiology Laboratory were nasopharyngeal and oropharyngeal swabs and placed in universal transport media (UTM, 350C, Copan Diagnostics, Murrieta, CA, USA). Samples obtained from the California Department of Public Health (CDPH) and SCDPH were nasopharyngeal and oropharyngeal swabs.

### Diagnostic Testing of SARS-CoV2

Quantitative PCR testing for SARS-CoV2 was performed by the CDC, California Department of Public Health, or the UCSF Clinical Microbiology Laboratory. At UCSF, samples were extracted on a Magna Pure 24 Viral Kit (Roche Diagnostics, Indianapolis, USA) or a EZ1 Viral Mini Kit (Qiagen, Hillden, Germany). At the California Department of Public Health, samples were extracted using Qiagen DSP Viral RNA mini kit with carrier RNA added (Qiagen). The one-step RT-PCR used the 2019-nCoV CDC qPCR Probe Assay targets N1 and N2, and the 2019-nCoV N Positive Control plasmid (Integrated DNA Technologies Inc. IDT, Coralville, USA) as a positive control. The RNA extracts were added to an amplification reaction mix (4X TaqPath 1-step RT-PCR master mix (Life Technologies, Carlsbad, USA), 1.5 ul primer (N1, N2, or RNase P), 12.5ul nuclease-free water, and 1ul RNA) and one-step RT-PCR was performed (25°C for 2 mins, reverse transcription at 50°C for 15 mins, 95°C for 2 mins, followed by 45 cycles PCR with denaturing at 95°C for 3s and extension at 55°C for 30s). The cut-off for a confirmed positive sample was determined to be at a Ct value of 40 cycles, with both targets of N1 and N2 necessarily being classified as positive by quantitative RT-PCR, except for a positive RNase P as an internal quality control.

### Whole Genome Sequencing of SARS-CoV2

The extracted RNA described above was converted to cDNA using MSSPE method (Metagenomic Sequencing with Spiked Primer Enrichment) as previously described (*15*). The custom designed SARS-CoV-2 primers (13-nucleotide in length, IDT Technologies) were constructed using 30 early SARS-CoV-2 genome references in the National Instiutes of Health (NIH) GenBank database, and were subsequently added to the reverse transcription process **(Table S1)**. The cDNA eluate was made into sequencing libraries through tagmentation (Nextera XT, Illumina), and were individually barcoded in preparation for Illumina sequencing. The samples were then bead-washed with AMPure XP beads (Beckman Coulter, Brea, USA) at 0.92X (23uL). The barcoded library then directly proceeds into a library recovery step. 5uL of the library is added to an amplification reaction mix (10uL Phusion 5X Buffer (ThermoFisher Scientific, Waltham, USA), 2.5uL of 10uM forward and reverse general primers, 1uL of 12.5mM dNTPs, 0.5uL Phusion enzyme (ThermoFisher Scientific, Waltham, USA), 31uL nuclease-free water) for 14 cycles, with cycling parameters as previously described (*22*). The amplicons were again purified using 0.9X volume of AMPure XP beads. At this point, the purified PCR product can be analyzed by gel electrophoresis to check library size.The eluted library was quantified using a Qubit fluorometer. Sequencing libraries were then sequenced on MiSeq, Nextseq, or HiSeq 1500 (Illumina Inc., San Diego, USA) as 1×150 single-end or 2×150 paired-end reads.

### Genome assembly and consensus generation

Raw reads were first screened via BLASTn (BLAST+ package 2.9.0) (*23*) for alignment to reference strain NC_045512. They were then aligned to the reference with LASTZ version 1.04.03. SARS-CoV-2 reads were trimmed using Geneious version 11.1.3 by removal of 13 nucleotides (nt) (the length of the MSSPE primer) and low-quality reads from the ends, followed by removal of duplicate reads. Trimmed reads were mapped to reference genome NC_045512 in Geneious with no gaps allowed and a maximum of 5% mismatches per read. The assembled contig was then manually annotated, and a consensus genome was generated using a majority threshold criterion.

### Phylogenetic analysis and genomic comparison

Sequences were aligned using MAFFT v7.427 (*24*) under default settings and multiple sequence alignments were manually corrected. Phylogenetic trees were constructed in PhyML v3.3 (*25*) under an HKY+Γ_4_ substitution model (*26, 27*). The location of SNPs and gaps were extracted from alignments using a custom Python script and visualized using a custom R-script.

### Data Sharing

The assembled SARS-CoV-2 genomes in this study were uploaded to GISAID (*28*–*29*) as FASTA files, and can be visualized on a continually updated phylogenetic tree on NextStrain (*30*). Submission of the genomes and raw sequence data to NIH GenBank and Sequence Read Archive (SRA) is pending.

## Data Availability

The assembled SARS-CoV-2 genomes in this study were uploaded to GISAID (28-29) as FASTA files, and can be visualized on a continually updated phylogenetic tree on NextStrain (30). Submission of the genomes and raw sequence data to NIH GenBank and Sequence Read Archive (SRA) is pending

https://www.gisaid.org/epiflu-applications/next-hcov-19-app/

## Acknowledgments

This work was funded by NIH grants R33-AI129455 (CYC) from the National Institute of Allergy and Infectious Diseases, the Charles and Helen Schwab Foundation (CYC), K08-CA230156 (WG), and the Burroughs-Wellcome CAMS Award (WG). We acknowledge the help and advice of James T. Lee at the US CDC.

## Author Contributions

CYC conceived, designed, and supervised the study. AG, AS-G, CW, CYP, GY, HG, VS, and XD performed experiments. CYC and SF assembled and curated the viral genomes. CYC, WG and XD analyzed data. CYC, EH, KZ, SM, and WG collected patient samples at UCSF. DM and GA analyzed genomic and epidemiologic data. TB, AG, PR, LMS, MF, HYC, JS, and KRJ collected, assembled, and provided viral genome data from Washington and contributed to the phylogenetic analysis. CA, KG, MZ, ES, and KGA provided viral genome data from Southern California. CP, JTL, JZ, ST, and YL sequenced and analyzed viral genomes at the CDC. LDP, NF and OP performed phylogenetic analysis of genomes. AY, BTM, BB, CC, DAW, EV, MP, MW, OK, SC, SM collected samples, extracted the viral RNA and/or provided epidemiology data from counties in California. CYC, WG, and XD wrote the manuscript. CYC, XD, SF, CW, DAW, LDP, OP, and WG edited the manuscript. All authors read the manuscript and agree to its contents.

## Disclosures

CYC is the director of the UCSF-Abbott Viral Diagnostics and Discovery Center (VDDC) and receives research support funding from Abbott Laboratories. CYC and XD are inventors on a patent application on the MSSPE method titled “Spiked Primer Design for Targeted Enrichment of Metagenomic Libraries” (US Application No. 62/667,344, filed 05/04/2018 by University of California, San Francisco). HYC is a consultant for Merck and GlaxoSmithKline, and receives research funding from Sanofi-Pasteur, Ellume and Cepheid, unrelated to this work. All other authors have no conflicts to declare.

The opinions expressed by the authors contributing to this journal do not necessarily reflect the opinions of the Centers for Disease Control and Prevention or the institutions with which the authors are affiliated.

## Supplementary Material

**Table S1: (Excel format) Custom primer sequences for the MSSPE (Metagenomic Sequencing with Spiked Primer Enrichment) method**

**Table S2 (Excel format). Metadata for 29 COVID-19 samples and corresponding genome sequencing metrics**

